# Understanding risk stratification of chronic kidney disease: qualitative study in primary care

**DOI:** 10.64898/2025.12.16.25342377

**Authors:** Kristin Veighey, Emma Teasdale, Michelle Myall, Kate Henaghan-Sykes, Tom Blakeman, Kinda Ibrahim, Bhargav Raut, Hazel Everitt, Simon DS Fraser

**Affiliations:** Primary Care Research Centre, University of Southampton, Southampton, UK; National Institute for Health and Care Research (NIHR) Greater Manchester Patient Safety Research Collaboration (GM PSRC), The University of Manchester, Manchester, UK; National Institute for Health and Care Research (NIHR) Manchester School for Primary care Research, The University of Manchester, Manchester, UK; National Institute for Health and Care Research (NIHR) Applied Health Collaboration (ARC) Wessex, Southampton, UK

**Keywords:** Chronic Kidney Disease, Risk stratification, Communication, Primary Care, Qualitative research, Prevention

## Abstract

**Background:** Chronic kidney disease (CKD) affects approximately 10% of adults in England. <1% progress to end stage-kidney disease (ESKD), significantly impacting health-related quality of life with high healthcare costs. CKD is associated with cardiovascular disease risk. New therapies to improve outcomes highlight the need for effective risk stratification.

**Aim:** To explore primary care teams’ views and experiences of CKD risk stratification.

**Design and Setting:** Qualitative interview and focus group study with GP practices in South England, South London and Yorkshire.

**Method:** 26 semi-structured interviews with GPs, pharmacists and practice nurses, and 4 focus groups (31 participants) with practice teams including clinical, administration and management staff, February 2024 to January 2025 across 20 practices, informed by normalisation process theory (NPT), using thematic analysis.

**Results:** We identified 4 key themes: 1) Awareness of diagnostic criteria and risk stratification tools, 2) Value of coding CKD and discussion of risk, 3) Barriers to CKD risk stratification, 4) Improving CKD risk stratification.

Despite universal awareness of diagnostic criteria, there was low awareness of risk stratification tools. Coding was perceived as valuable for health professionals but not for patients. Concerns included increasing patient anxiety and over-medicalisation. Time pressures and lack of incentivisation were perceived as key barriers. Improved healthcare professional education, guidelines, pathways and technology/automation were highlighted as areas of potential improvement.

**Conclusion:** Primary care awareness of CKD is high, but workload, time pressures, and concerns regarding patient anxiety and over-medicalisation contribute to incomplete risk stratification. Refining risk stratification procedures and effective patient communication could improve care.

## Introduction

Chronic kidney disease (CKD) affects around 10% of adults in England (1, 2). Approximately 1%, (around 8,254 UK adults in 2022 (3) will progress to end-stage kidney disease (ESKD) requiring kidney replacement therapy (KRT). It has significant health-related quality of life and economic impacts. NHS treatment costs for CKD are over £6 billion per year, >3% of the overall budget, with dialysis treatment costing around £34,000 per patient each year (4). The World Health Organisation (WHO) designated kidney disease a global health emergency in 2025 (5).

CKD is defined by the Kidney Disease Improving Global Outcomes (KDIGO) consortium as a persistent (over 3 months) structural or functional abnormality in kidney function (1, 6). Kidney disease is risk stratified using blood tests for kidney function (estimated glomerular filtration rate, eGFR: G1 - normal or high, to G5 - kidney failure) (6,7), and urine tests for kidney damage (urine albumin:creatinine ratio, uACR: A1 – normal to mildly increased to A3 – severely increased) (7). As eGFR falls, and albuminuria increases, both cardiovascular disease and ESKD risk increase. The Kidney Failure Risk Equation (KFRE) uses age, sex, eGFR and uACR to provide a 5-year risk estimate of ESKD – i.e. renal risk - in those with CKD Stage 3-5. The UK National Institute for Health & Care Excellence (NICE) advocates those with a KFRE score > 5% should be considered for referral to secondary care (8). Both KDIGO staging and the KFRE require both eGFR and uACR, however since withdrawal as a Quality and Outcomes (QoF) indicator in 2014, uACR is only completed in around 50% of patients with CKD (9). People living with CKD are overall at greater risk of cardiovascular disease than of ESKD, with uACR predicting cardiovascular risk (10).

Optimal CKD management entails informed communication with patients with risk stratification to explain disease severity and its broader implications. Many patients with CKD have multi-morbidity. Frailty, diabetes, hypertension and obesity/metabolic syndrome can impact disease and progression.

This study explored the views and experiences of primary care professionals and the wider GP practice team regarding CKD, including factors underpinning implementation of CKD risk stratification and management in primary care and the complex mechanisms that influence how innovations are adopted, adapted, and sustained in practice.

## Methods

### Design

A qualitative study comprising semi-structured online interviews with healthcare professionals and focus groups with whole practice teams. recruited from Southern England (Hampshire, Isle of Wight and Dorset), South London and Yorkshire, to reflect rural, coastal, and inner-city populations.

The gender-diverse team comprised two research fellows with experience of conducting qualitative research, a GP clinical academic registrar, a GP registrar, two clinical academic GPs a public health academic, an academic pharmacist, an implementation science expert, and a patient and public involvement (PPI) contributor. Core research team members held monthly meetings to review study progress, review interview data, and collaborate on data analysis. Additional separate analysis meetings were held with the PPI contributor.

This study is reported in accordance with the Standards for Reporting Qualitative Research (11).

### Participants and recruitment

GP practices were invited to participate via the National Institute for Health Research (NIHR) Clinical Research Networks (CRNs, now Regional Research Delivery Networks (RRDNs)) in Wessex, South London and Yorkshire. Study information invited primary care professionals to take part in an individual interview and/or the whole practice to take part in a focus group. Focus groups were drawn from different areas i.e. rural, coastal, inner city (Dorset, Hampshire and the Isle of Wight, South London and Leeds) to include practices with a range of patient demographics, including age, ethnic diversity and levels of deprivation.

Potential participants received an invitation pack containing study information and consent form and could choose to participate online or in person. In addition to written informed consent, interviewers obtained verbal consent before starting each interview or focus group.

### Data collection

Separate interview and focus group topic guides drew on the existing literature and input from our patient collaborator. Topic guides (Supplementary Box 1) were informed by key domains from normalisation process theory (NPT) (12), which provides a framework for identifying factors and processes likely to hinder or enable widespread implementation of new practices. They captured data on value and purpose (Coherence), participant’s engagement (Cognitive Participation), the work involved (Collective Action) and appraisal work undertaken (Reflexive Monitoring). Interviews, and online focus groups, were audio-recorded using Microsoft Teams. In-person focus groups were recorded using an audio recorder. Field notes were taken to capture the interviewers’ impressions and reflections, and any aspects not captured by the recorder.

Interviews lasted 25 to 64 minutes (mean 40 minutes). Focus groups lasted 27 and 38 minutes (mean 34 minutes) and were conducted during existing practice meeting timeslots. On finishing each interview or focus group, interviewees were offered a copy of their transcript and study findings, when available. Data collection stopped on reaching thematic saturation (that is, when themes were rich, well developed, and understood) and a rigorous, credible analysis in relation to our aims had been achieved.

### Data analysis

All recordings were transcribed verbatim by a professional transcription company and any identifiable data removed. Thematic analysis (13) was used to explore the data. Data collection and initial analyses proceeded iteratively (that is, coding started after the first few interviews and informed subsequent interviews). Interview and focus group transcripts were initially coded separately. Two lead authors (ET and KV) read the transcripts repeatedly to achieve data familiarisation and then applied line by line coding. Codes were derived inductively from the data and grouped together to produce an initial coding frame by recognising meaningful repeated patterns and identifying key concepts in the data. Initial codes were reviewed and compared to identify any similarities and differences and a detailed coding manual created to ensure transparent and systematic coding. Interview and focus group data were compared and combined into overarching themes. Themes/subthemes were discussed and iteratively developed in collaboration with the research team to ensure diverse inferences and data interpretation. A negative case analysis was undertaken ensuring all data was carefully considered and reducing the chance of preferentially selecting data fitting any preconceptions. Although analysis was primarily inductive (data-driven), normalisation process theory (NPT) guided interpretation through input from two authors (KI and MM) who have significant experience in NPT. Themes and sub-themes were reviewed, reassigned and mapped against the four constructs of NPT. (Supplementary Box 2). NVivo (version 12) was used to manage data, implement and record coding.

## Results

January 2024 to January 2025, two researchers conducted 26 online interviews with GPs, pharmacists and nurses and 4 focus groups (three in-person and one online) with GP practice teams including healthcare professionals, social prescribers, health coaches, administration and management staff (31 participants in total). Participant characteristics in Table 1.

**Table 1:** Participant Characteristics. *7 focus group participants did not provide demographic data

| Baseline Characteristics | Interviewees<br>(n=26) n (%) | Focus group members<br>(n=24)* n (%) |
| --- | --- | --- |
| **Age, range (mean)** | 30-64 (45) | 29-52 (45) |
| **Gender** |  |  |
| Female | 19 (73) | 23 (96) |
| Male | 7 (27) | 1 (4) |
| **Ethnicity** |  |  |
| Arab | 2 (8) | 0 (0) |
| Asian/Asian British | 4 (15) | 1 (4) |
| Black/African/Caribbean/Black British | 2 (8) | 1 (4) |
| White | 17 (65) | 21 (88) |
| Mixed / Multiple | 1 (4) | 1 (4) |
| **Job Role** |  |  |
| GP | 21 (81) | 6 (25) |
| Nurse/Nurse Practitioner | 1 (4) | 3 (13) |
| Pharmacist | 4 (15) | 4 (17) |
| Administration/Clerical/Manager | NA | 7 (30) |
| Other (social prescribers, care co-ordinators, HCA etc.) | NA | 4 (17) |
| **Length of time since CCT** |  |  |
| Less than 2 years | 0 (0) | 1(6) |
| 2-5 years | 1 (4) | 5(29) |
| 6-9 years | 7 (27) | 1 (6) |
| 10+ years | 18 (69) | 10 (59) |
| **Length of time practice** |  |  |
| Less than 2 years | 2 (8) | 4 (17) |
| 2-5 years | 6 (23) | 12 (50) |
| 6-9 years | 8 (31) | 4 (17) |
| 10+ years | 10 (38) | 4 (17) |
| **Region** |  |  |
| Wessex | 17 (65) | 3 |
| Yorkshire | 6 (23) | 1 |
| South London | 3 (12) | 0 |

### Key themes

We identified 4 key themes:1) Awareness of diagnostic criteria and risk stratification tools, 2) Value of coding CKD and discussion of risk, 3) Barriers to CKD risk stratification, 4) Improving CKD risk stratification (Figure 1). Themes are presented below with selected illustrative anonymised quotes (FG = focus group participants, P = interview participants)

**Figure 1:**
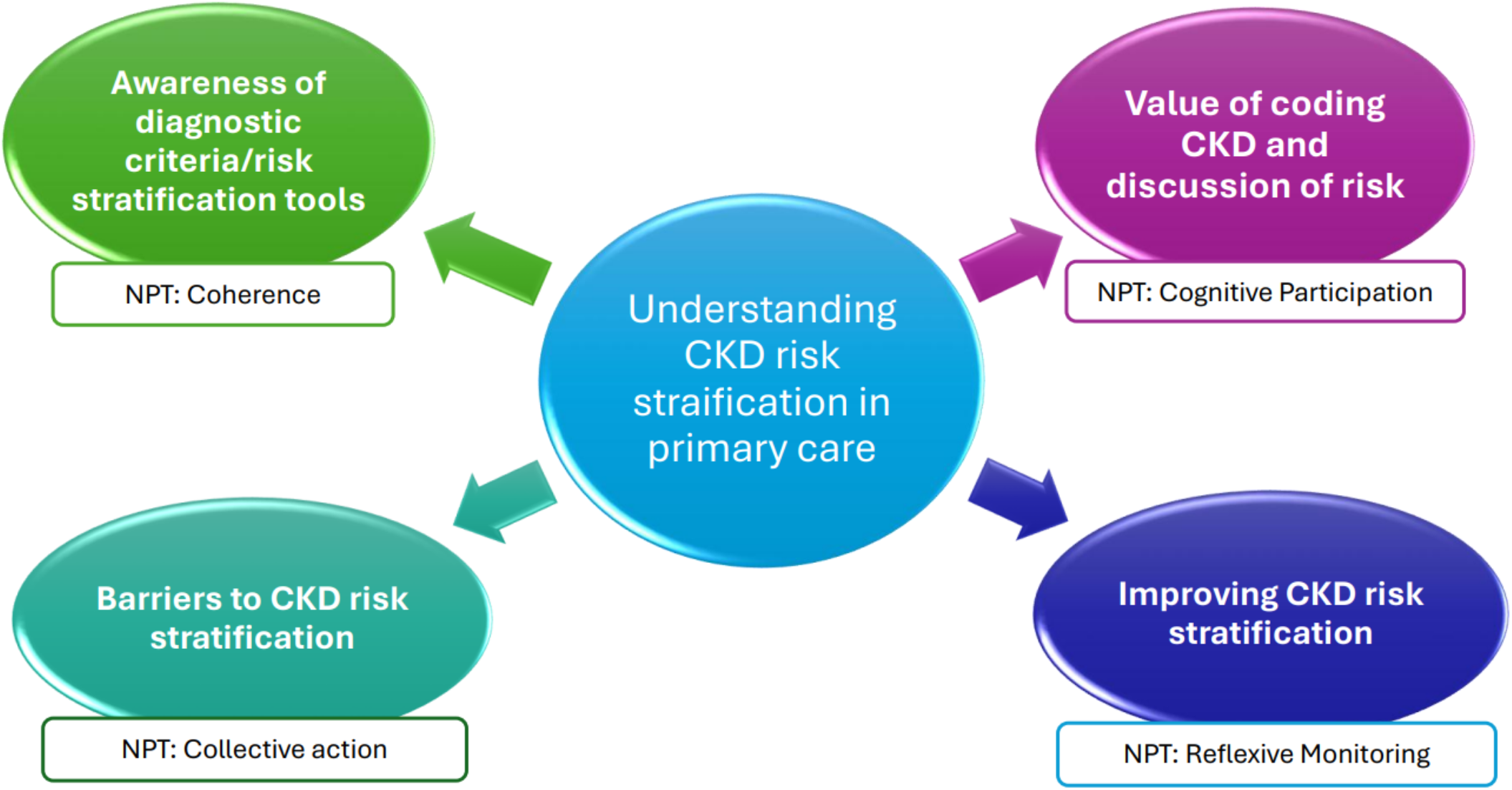
Key themes and related NPT constructs

**Figure 2:**
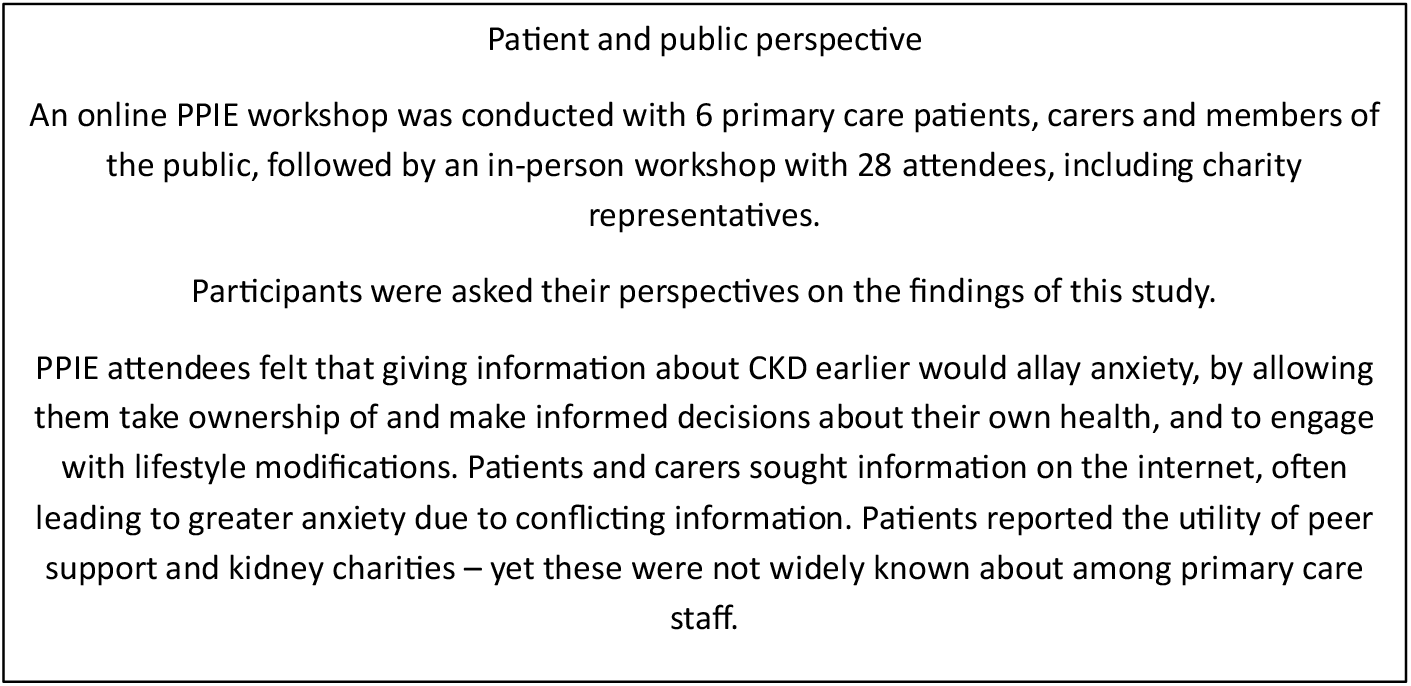
Patient and public perspectives of the research findings

#### Awareness of diagnostic criteria and risk stratification tools

Across the multidisciplinary primary care team, participants demonstrated a shared and consistent understanding of the diagnostic process for chronic kidney disease (CKD). Interviewees described use of diagnostic blood and urine tests (eGFR and ACR) in keeping with guidelines, suggesting familiarity. Participants differentiated CKD from other chronic conditions through their recognition of its typically asymptomatic nature and the incidental way it is often identified. This differentiation (a key subcomponent of ‘Coherence’ within the NPT Framework) highlights that while CKD is conceptually understood, it is often normalised as part of routine monitoring rather than as an active diagnostic focus.

> *“So generally, it’s opportunistic. We have our chronic disease patients that are invited*…*to do a urinary ACR as well. So they’ll be doing those annually…but generally it’s opportunistic*.*”* (FG3, Female Pharmacist)

Analysis of participants’ accounts suggested that CKD diagnostic activities sit within a broader framework of chronic disease management, but the process is largely driven by existing systems and opportunistic testing rather than proactive identification.

In contrast to the strong shared understanding of diagnostic processes, there was limited collective understanding (Coherence) around CKD risk stratification. Few participants reported using formal risk stratification tools, and awareness of existing instruments—such as the Kidney Failure Risk Equation (KFRE) or the KDIGO risk grid—was low.

While participants recognised the potential utility of such tools, their comments reflected uncertainty about how they fit within existing workflows and clinical priorities.

> *“I can see how referring those patients early is important. Perhaps we can prevent some progression if we were risk stratifying a bit better those patients*.*”* (P16, Female GP)

#### Value of coding CKD and discussion of risk

Diagnosing and coding CKD in clinical systems was typically perceived as important and valuable to health professionals, mainly to ensure safe prescribing, but consistently felt to be less useful for patients.

> *“When you’re coming to presc*ribing as a nurse practitioner, if I can see a diagnosis of CKD, I’m obviously going to be far more wary about what I’m prescribing…So I think for healthcare professionals, really important. For patients, not so much.” (P3, Nurse Practitioner)

This led to challenges around forming a clear value and purpose ‘Coherence’, in securing engagement ‘Cognitive Participation’ and willingness to undertake the work involved ‘Collective Action’. Participants from across the multidisciplinary primary care team reflected on the potential to cause patient anxiety by coding/diagnosing early-stage CKD, especially in older people, and the risk of over-medicalising. Practitioners commonly described the language around CKD as challenging, easily leading to patient misunderstanding, anxiety and concerns. They highlighted a lack of societal awareness around kidney health, and the common difficulties of informing patients of historical CKD coding of which they were unaware.

> *“It’s generating workload, generating disease, not disease mongering, but disease pathologizing of patients with a scary title. Lots of our patients, lots of anxiety out there, it’s unrecognised. Lots of health anxiety. Are we potentially feeding into that and causing some harm with marginal benefits? So, I guess that’s a potential barrier or driver for us not being not being as keen as we could be to code*.*”* (P22, Female GP)

However, some participants acknowledged the need for risk stratification to optimise care and viewed coding, giving a diagnosis of CKD and discussing risk of progression as an opportunity for health promotion.

“*When I started in general practice it was like, ‘Oh, your kidney function is not so good, don’t worry about it,’ it’s not like that anymore, obviously, or, ‘It’s just age related,’* …*Whereas now, I think we’re much more actually, we can do something about it, we can modify your risks, we can ensure that okay, your kidney function may not be as good but actually, we can try and prevent you having some of the complications associated with that*.*” (P16, Female GP)*

#### Barriers to CKD risk stratification

Time pressures in general practice and lack of incentivisation were key challenges to diagnosing and risk stratifying CKD. Across accounts, participants described described finding coding CKD challenging as it was often done during reviewing results of patients they may not know personally. This contributed to challenges related to engagement in a coherent plan for prioritisation, reduced engagement ‘Cognitive participation’ and influenced ‘buy-in’ ‘Collective Action’ which could limit meaningful patient outcomes. Participants articulated the complexity of what is required in each CKD consultation as a challenge.

> *“Just how time-consuming it is, really. Even on an individual basis, if you’re seeing someone for a kidney disease, there’s so much to go through, it is just quite time-consuming*…*the consultation is quite time-consuming*.*” (P8, Female GP)*

Lack of incentivisation of coding, diagnosing and risk stratifying CKD was another common perceived barrier to good care. Participants acknowledged that a lack of incentivisation fuels the concept that CKD is not as important as other incentivised diseases. The large number of patients with CKD, the lack of processes to risk stratify which patients to prioritise, opportunity within routine work, and cost-effectiveness concerns were highlighted. Lack of a clear risk stratification process led to operational work during clinical encounters being fragmented. Without guidance on which patients to prioritise, clinicians faced challenges integrating risk stratification into routine consultations.

> *“I mean kidneys literally are the forgotten organ as far as I’m concerned. There is no QOF indicators, there’s nothing hanging on it*…*my organisation has got 47,000 patients. When you start running things like searches on how do we stratify these patients or find the most important ones that we can start making a difference for, the numbers are humungous*.*”* (P9, Female GP)

Patient health literacy and ability/willingness to engage with suggested advice and management plans following a CKD diagnosis was highlighted in the focus groups. Healthcare professionals expressed concerns some individuals may not engage with lifestyle measures or comply with medications for diabetes or blood pressure particularly when asymptomatic. This was a particular concern for those already known to be marginalised by existing models of healthcare, such as those living in areas of high deprivation, on benefits, with other long-term conditions, and minority ethnic groups – especially where there was already cultural hesitancy or mistrust of standard healthcare systems.

> *“That’s not everyone at all, but it is a significant issue in an area of high deprivation because patients just don’t… It’s very low down on their list of priorities to know what a kidney is, or when their kidneys are not working. They’ve got so many other things that they worry about more, like where are they going to sleep, how are they going to pay for their heating, that kind of thing*.*” (FG1, Female GP)*

#### Improving CKD risk stratification

Mechanisms suggested to improve CKD care included: improved pathways, guidelines, education, testing processes, integrated technology/automation, incentivisation and community-based clinics. Research studies and incentivisation for coding and risk stratifying (e.g. established payments for prescribing statins for CKD) were described as an impetus for system wide searches and increased coding.

Improved technology (such as point-of-care testing), automated diagnostic/coding support included within results (for healthcare professionals) and associated signposting (for patients) was suggested to support coding and risk stratification, as ways of contextual integration within collective action.

> *“I have no idea why EMIS can’t do coding automatically and pull down data from the hospital system in the same fashion. So, it’s really frustrating that we have to go into the hospital for blood tests to pull them down to then be able to compare because invariably, a lot of those will be missed. So, I think that it seems silly that IT lets you down, really, in primary care still. It shouldn’t be really done by doctors or anyone else. It should all be automatic, so I think, yes, that the technology needs to be better definitely… I think AI technology, actually, in this scenario, I think it would be a huge benefit for patients*.*” (P10, Female GP)*

Creating dedicated community CKD clinics and specialist CKD health professionals was commonly seen as a mechanism to provide additional resource allowing time for effective discussion of risk and encourage improved risk stratification and CKD care.

“*I think if we had*…*chronic kidney disease clinics, because we’re seeing more and more*…*we could get patients to think that, okay, chronic kidney disease is quite common, or it doesn’t shock them, and maybe make them more aware that chronic kidney disease is something that we see quite regularly in primary care and maybe we can deal with it via primary care*.*” (p19, Female Pharmacist)*

Education programmes and tailored patient resources, as for other long-term conditions such as diabetes, were seen as effective ways to deliver reliable information efficiently and support discussion of risk.

> *“For our pre-diabetes patients, there’s a diabetes education programme they can do, there’s some information they can easily access about that. I’m not aware, and I could be wrong, that there’s anything really for CKD. If we could have somewhere we could easily signpost the patients and have a conversation with them but not having - if you don’t have the time to have an in-depth conversation, having somewhere to signpost them that they could go for that*.*” (P16, Female GP)*

> *“having culturally appropriate leaflets or information or even videos of people with lived experience of CKD of similar backgrounds, I think sometimes that’s more powerful than a conversation with a doctor*.*” (P25, Female GP)*

## Discussion

### Summary

This qualitative research explored primary care healthcare professionals’ views on risk stratification for patients with CKD, through individual practitioner interviews and whole practice team focus groups. The application of NPT as the theoretical lens enabled identification and exploration of the variety of work and processes needed to successfully undertake CKD risk stratification in primary care.

Findings highlighted an absence of systematic approaches to CKD risk stratification. NICE diagnostic criteria were widely recognised, but CKD diagnosis was often opportunistic, and awareness of risk stratification tools was low. From the perspective of NPT these findings suggest limited *coherence* — while clinicians understood CKD as a clinical entity, its proactive identification and management were not fully conceptualised as routine work (14, 15). Coding is known to improve care in both UK (16) and European (17) cohort studies, however whilst our participants valued diagnostic coding for professional purposes such as safe prescribing, it was perceived as less meaningful for patients, particularly where concerns about inducing anxiety were prominent. These tensions indicate challenges in *cognitive participation* and *collective action*, as professionals negotiated competing priorities between patient-centred communication, avoiding over-medicalisation and safety-driven administrative tasks (18).

Practical difficulties of informing patients at the point of diagnosis, coupled with time constraints and prioritisation of coding, further illustrate incomplete *normalisation* of planned, relational care processes. Knowledge gaps among reception staff and limited awareness of community resources were illustrative of limited *collective action* at the micro level, where front-line teams lacked capacity to support patients effectively. Similarly, nursing teams acknowledged potential roles in promoting engagement with monitoring (e.g., uACR) but reported inconsistent enactment, reflecting insufficient *reffexive monitoring* of team contributions.

We found organisational barriers, including lack of incentivisation and time, despite recognition of the alignment of CKD risk stratification with the NHS 10-Year Plan (19). Participants proposed leveraging existing frameworks for hypertension and diabetes management and involving specialist nurses and pharmacists, suggesting attempts to embed CKD work within established systems—a key NPT mechanism for sustaining practice (20). However, these adaptations were contingent on resource allocation and organisational support to ensure collective action at a meso level.

Policy ambitions to improve CKD prevention and management were evident, yet implementation was impacted by longstanding systemic inequities of access and engagement and workforce pressures. Persistent disparities in engagement among patients most at risk of CKD, e.g. those with low socioeconomic status, poor health literacy, and minoritised backgrounds reflect health inequalities (NHS England, Core20PLUS5) are compounded by limited availability of tailored resources. These findings emphasise the need for coordinated strategies that address structural determinants of health and provide adequate resourcing for primary care teams.

Our findings demonstrate that while CKD risk stratification was recognised as clinically important, its integration into routine practice was hindered by gaps in coherence, cognitive participation, and collective action at the individual and organisational levels. An approach to improve Testing, Triage (risk stratification) and Treating holistically has the potential to improve outcomes.

### Strengths and limitations

This study explored the views of primary care healthcare professionals towards the rapidly changing landscape of CKD guidance and management adding valuable new insights into individual and system level factors effecting care. Most people CKD are cared for in primary care whereas much of the existing literature centres around people within secondary care clinics who have more severe CKD and established CKD diagnoses.

The study recruited in a few limited regions and interviews were likely to have taken place with more engaged practitioners and practices, which may limit generalisability. There may also be desirability bias in responses to interviews and focus groups.

This study interviewed only health care staff as there is a greater published evidence base for patients with CKD. However, previous patient work has also been mostly focussed on patients managed in secondary care so the views of people with less severe CKD in primary care settings needs to be explored further.

### Comparison with existing literature

A qualitative study in 2012 (21) explored processes underpinning the implementation of CKD management in primary care, through interviews with GPs and practice nurses. A predominant theme was anxiety about disclosure of CKD, especially in more elderly patients (21), in keeping with findings from our study. A further qualitative study in 2015 explored GPs views of managing advanced CKD and again expressed a wish for better guidance around managing older patients with advanced CKD and co-morbidities (22). The impact of non-disclosure of CKD in primary care on patient ability to self-manage has also been highlighted (23). A narrative review in 2024 described the UK primary care perspective of barriers and enablers to effective kidney care but did not directly seek to use qualitative research to explore these views (24).

Since 2012, national endeavours such as the NHS England/UK Kidney Association Think Kidneys/Transforming Participation in Chronic Kidney Disease programme (25) have aimed to raise the profile of kidney disease. The NHS England Renal Service Transformation Programme (RSTP) concluded in 2024 with the creation of 8 regional secondary care led renal clinical networks, with the goal of improving integrated care. However, this study highlights that while awareness of CKD and its importance is widespread in primary care, there are significant challenges in implementing risk stratification, and hence equitable access of novel evidence-based treatments.

### Implications for research and/or practice

The study findings highlight that Integrated care pathways for CKD are urgently needed, alongside a public awareness campaign and healthcare professional education. Integration of laboratory systems, and the visibility and articulation of risk-based information for patients at the time of coding within resources such as the NHS App should be co-produced, refined and evaluated.

Clinicians reported concern about causing anxiety to patients, yet PPIE contributors stressed the importance of even earlier disclosure of a CKD diagnosis to aid education, lifestyle changes and patient empowerment to self-manage. Training in consultation skills to support conversations with patients around CKD risk could reduce HCP anxieties around these consultations.

This study reinforces the need for better communication with patients and the public around CKD risk, which is now urgent given the advent of effective evidence-based therapies to reduce both cardiovascular and renal risk. Emerging evidence from large cohort studies reinforces that this proactive approach improves both kidney and cardiovascular outcomes and reduces environmental burden from CKD (26).

Due to the high prevalence of CKD, and a clinical trial evidence base which recruited mainly younger secondary care patients, further work is needed to confirm cost effectiveness in the primary care setting, and confirm the risk:benefit ratio of treatments in older population cohorts.

## Data Availability

All data produced in the present study are available upon reasonable request to the authors

## Additional information

## Ethical approval

University of Southampton Ethics and Research Governance Online (ERGO) reference 81055 and Health Research Authority Integrated Research Application System (IRAS) Health Regulatory Authority (HRA) Reference Number 330328

## Acknowledgements

Mr Charles Pickering, patient contributor

